# Needlestick and Sharp Injuries Prevalence and Hepatitis B Vaccination Among Healthcare Workers: A cross sectional study in Six District Hospitals (Yaounde, Cameroon)

**DOI:** 10.1101/2023.04.14.23288599

**Authors:** Fabrice Zobel Lekeumo Cheuyem, Emilia Enjema Lyonga, Hortense Gonsu Kamga, François – Xavier Mbopi – Keou, Innocent Takougang

**Author notes:** Corresponding author. Address: Innocent TAKOUGANG, Department of Public Health, Faculty of Medicine and Biomedical Sciences, The University of Yaounde I, PO Box: 8526, Yaounde, Cameroon.

## Abstract

**Introduction:** Accidental exposure to blood and body fluids (AEB) in the workplace account for 40 % of contamination by hepatis B virus (HBV) and 2 – 3 % by HIV among healthcare workers (HCW). Developing countries are most affected. The present study sought to determine the prevalence of percutaneous injury and hepatitis B vaccination coverage among HCW.

**Methods:** A cross-sectional study was carried out from January to April 2022 in six district hospitals in Yaounde using a self - administered questionnaire. Out of the 279 HCW who were solicited, 217 returned completed questionnaires.

**Results:** More than half of HCW reported an AEB in the last 12 months (53,9 %). The prevalence of AEB varied among hospitals with the Nkolndongo DH reporting the highest prevalence (51.6 %). Healthcare workers were unvaccinated (53 %) or partially vaccinated against HBV (13,2 %); only one third were fully vaccinated (33,9 %). The lowest compliance with vaccination was observed among hygiene personnel (90 %). The high cost of the vaccine was the main reported reason for non - compliance (39 %).

**Conclusion:** There is an urgent need to set up a monitoring system for the implementation of infection control and prevention in District Hospitals in Cameroon in order to raise awareness of AEB burden among healthcare workers and improve accessibility to HBV vaccine.

## Introduction

Healthcare workers experience needlestick, sharp cuts and splashes with biological fluids while providing health care [1,2]. It is estimated that three million HCWs are exposed each year resulting in approximately 170 000 HIV, two million viral hepatitis B (HBV), and 0,9 million viral hepatitis C (HCV) infection [3,4]. Percutaneous accidental exposures to blood are injuries caused by needle, sharp object and broken items that are contaminated with body fluids [5]. Blood exposure accidents affect developing countries most, where the prevalence of AIDS, HBV infections are highest [6]. A systematic review conducted in 21 African countries found a high prevalence of occupational exposure to body fluids; two thirds of HCW experience at least one exposure during their career [7]. Public and in private health facilities are equally affected [8]

Occupational exposure to blood is associated with work environment and institutional determinants, attitudes of HCW and patient related factors [9–13]. Work environment factors include professional stress, work load, availability of health and security information and training, clinical service (surgical ward), professional experience and availability of personal protective equipment (PPI) supplies [9,10,13,14].

Healthcare workers related conditions such as age, gender, professional status, hepatitis B vaccination status, recapping needle can influence the occurrence of accidental exposure to body fluid [9,11]. Besides, poor adherence to infection control and prevention measures such as handwashing and systematic use of PPI during healthcare can also significantly drive accidental of exposure to body fluid among healthcare workers. Moreover, poor patient compliance during healthcare, especially the sudden movement during blood sampling is associated to occupational exposure to blood and body fluids [10,12,13].

The risk of exposure infection transmission from blood varies according to different epidemiological and contextual factors. Epidemiological components including infection prevalence in the general population. Among contextual factors, we distinguish the depth of the injury, nature of the accident (prick, cut, projection), absence of PPI (googles, gloves), healthcare worker vaccination status, nature of the fluid associated to the exposure, time between exposure and consultation and serological and clinical status of the source patient [4,15].

Healthcare workers are one of the most vulnerable groups, with up to four times greater risk of contracting the infection than the general population [16]. Moreover, Sub-Saharan African countries have a high prevalence of blood borne infections in the general population [7,17]. Although Cameroon introduced the anti - HBV vaccine in their Expanded Program of Immunization in 2005, this program does not cover HCW [18]. Besides, there is a lack of awareness of the transmissibility of HBV among HCW [19]. The present study aimed to assess the level of exposure of HCW and viral hepatitis B vaccination status among HCW in Yaounde district hospitals (DH).

## Methods

### Study design & period

We conducted an institutional - based cross - sectional study in the six district hospitals of Yaounde from January to April, 2022.

### Setting

Yaounde, Cameroon’s capital, is host to a population of 3.2 million. It is the country’s second largest city [20]. The Cameroonian health system is organised around health districts. A district hospital is the first level of reference in the health pyramid responsible for providing primary health cares to the population [21]. The Yaounde DHs (Biyem - Assi, Cite - Verte, Djoungolo, Efoulan, Mvog Ada and Nkolndongo) cumulate nearly 400 health personnel, 330 beds with 153 543 consultations in 2020 [22].

### Participant

The study population consisted of workers who are in contact with patients and potentially exposed to body fluids. They were doctors, nurses, midwives, nursing assistants, laboratory technicians and cleaners.

### Sample size

An exhaustive sampling method was adopted in each clinical department, including all consenting personnel.

### Data collection

The study instrument was a structured self – administered questionnaire consisting of 17 questions covering sections related to socio - professional characteristics, experience of exposure to body fluids and hepatitis B vaccination status.

### Data processing and analysis

All filled questionnaires were entered and analysed using IBM SPSS Statistics (Statistical Package for Social Science) 2019 Version 26.0.0.0 software. The Chi-square (*X*^*2*^) test or Fisher’s exact test for proportions were used to compare proportions. Multivariate logistic regressions were used to assess the strength of the association between variables. A *p* - value < 0.05 was considered statistically significant.

## Results

Out of the 279 HCW contacted, 217 returned the completed questionnaire, representing 78 % response rate. Most of our study participants were female (81 %). Participants between 25 - 39 years were the most represented (73.7 %). They were mostly nurses (32.3 %) and laboratory technicians (21.2 %). Most participants had 7 - 10 years of professional experience (Table I and II).

**Table I.** Type of accidental exposure to blood among healthcare workers in Yaounde District Hospitals, April 2022 (*n =* 217)

| Type of exposure | Exposure ≤ 12 months | | | |
| --- | --- | --- | --- | --- |
|  | Count<br>(n) | Frequency<br>(%) | 95 % CI limits |  |
|  |  |  | Lower | Upper |
| Percutaneous injury | 64 | 29,5 | 23,5 | 36,0 |
| Splash | 101 | 46,5 | 39,8 | 53,4 |
| Exposure over the career |  |  |  |  |
| Percutaneous injury | 149 | 68,7 | 62,0 | 74,8 |
| Splash |  | Not measured |  |  |

**Table II.**
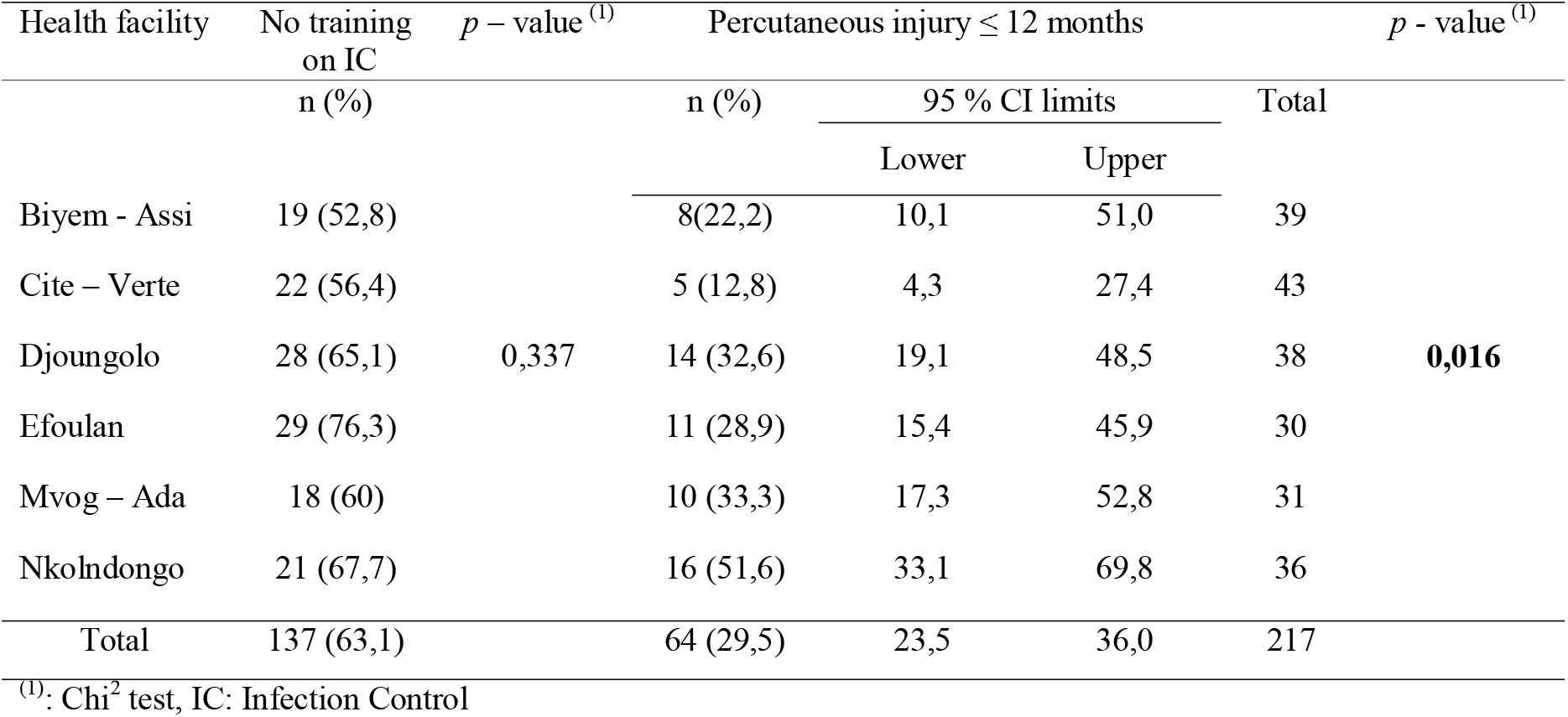
Experience of exposure to blood and training on infection control within Yaounde District Hospitals, April 2022

### Experience of needlestick injury

More than half of our participants had experience at least one AEB in the last 12 months (54 %). Most exposures resulted from percutaneous stings (29.5 %) and splashes (46.5%). More than two third of HCW declared having suffered an AEB over their career (69 %).

Over the last 12 months, the Nkolndongo DH witnessed the highest prevalence (51.6 %) followed by Mvog - ada DH (33.3 %). Cite - Verte DH had the lowest prevalence (12.8 %). Nkoldongo (67,7 %) and Efoulan DH (76,3 %) presented the highest proportions of HCW with no routine training on infection control and prevention (Table II).

There was no significant difference in exposure between sex (*p -* value = 0.849). Health personnel affected by percutaneous AEB were mainly nurses (42.9 %), students (33.3 %) and laboratory technicians/dentists (23.9 %). These differences were not statistically significant (*p -* value = 0.084) (Table III).

The surgery department recorded the most cases of percutaneous exposure to blood (47.1%) (Table IV).

**Table III.** Experience of exposure according to participants’ sociodemographic characteristics in Yaounde District Hospitals, April 2022 (*n =* 217)

| Characteristic | Count (n) | Frequency (%) | Total (100 %) | *p* - value |
| --- | --- | --- | --- | --- |
| **Gender** |  |  |  |  |
| Female | 51 | 29,0 | 176 | 0,849<sup>(1)</sup> |
| Male | 13 | 31,7 | 41 |  |
| **Age (year)** |  |  |  |  |
| 18 – 24 | 3 | 17,6 | 17 | 0,329<sup>(2)</sup> |
| 25 - 34 | 31 | 37,3 | 83 |  |
| 35 - 44 | 20 | 26,0 | 77 |  |
| 45 - 49 | 7 | 29,2 | 24 |  |
| 50 and + | 3 | 18,8 | 16 |  |
| **Marital status** |  |  |  |  |
| Single | 30 | 30,6 | 98 | 0,428<sup>(1)</sup> |
| Married | 34 | 28,6 | 119 |  |
| **Professional status** |  |  |  |  |
| Assistant nurse | 4 | 22,2 | 18 | 0,329<sup>(1)</sup> |
| Student | 4 | 33,3 | 12 |  |
| Nurse | 30 | 42,9 | 70 |  |
| Doctor | 10 | 27 | 37 |  |
| Hygiene technician | 2 | 10 | 18 |  |
| Midwife/Birth attendant | 3 | 21,4 | 14 |  |
| Laboratory/Dental Technician | 11 | 23,9 | 46 |  |

<sup>(1)</sup>: Chi<sup>2</sup> test, <sup>(2)</sup>: Fisher exact test

**Table IV.** Experience of exposure according to participants’ socio-professional characteristics in Yaounde District Hospitals April 2022 (*n =* 217)

| Unit | Count (n) | Frequency (%) | Total (100 %) | *p* - value |
| --- | --- | --- | --- | --- |
| Surgery | 16 | 47,1 | 34 | 0,084<sup>(1)</sup> |
| Hygiene and sanitation | 2 | 10 | 20 |  |
| Laboratory service | 13 | 25,5 | 51 |  |
| Obstetrics & Gynaecology | 10 | 32,3 | 31 |  |
| Internal medicine | 13 | 35,1 | 37 |  |
| Stomatology | 3 | 19,7 | 18 |  |
| Paediatrics | 7 | 26,9 | 26 |  |
| **Professional experience (year)** |  |  |  |  |
| 0 – 3 | 20 | 38,5 | 52 | 0,384<sup>(2)</sup> |
| 4 – 6 | 11 | 30,6 | 36 |  |
| 7 – 10 | 18 | 24,7 | 73 |  |
| 11 and + | 15 | 26,8 | 56 |  |

<sup>(1)</sup>: Fisher exact test; <sup>(2)</sup>: Chi<sup>2</sup> test

### Hepatitis B vaccination

More than half of our participants were unvaccinated (53 %) while one third were fully vaccinated (34 %) (Figure 1).

**Figure 1:**
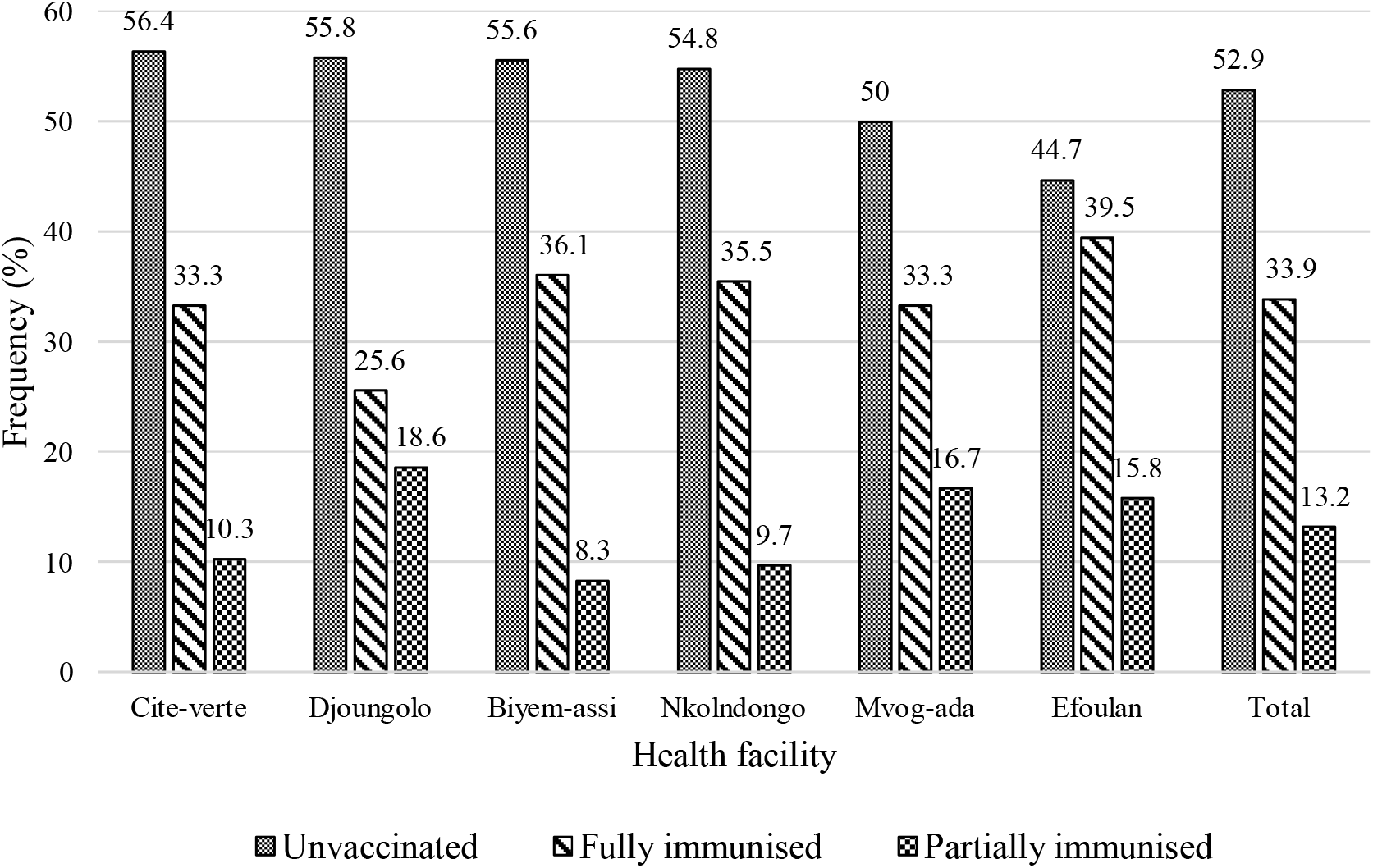
Compliance with hepatitis B virus vaccination among healthcare workers in Yaounde District Hospitals, April 2022 (*n =* 217)

Most hygiene personnel (90%) and paramedics (66.2%) were not completely vaccinated. Nearly two - third of participants (64.8 %) who had undergone percutaneous exposure to blood were not fully immunized against HBV (Table V).

**Table V.** Hepatitis B vaccination status according to socio – professional category and percutaneous exposure status among healthcare workers inYaounde District Hospitals, April 2022 (*n* = 217)

| Socio-professional category | HBV vaccination status<br>n (%) |  |  | Total | *p* - value |
| --- | --- | --- | --- | --- | --- |
|  | Unvaccinated | Partially | Fully |  |  |
| Medical | 15 (44,1) | 3 (8,8) | 16 (47,1) | 34 (100) | **0,041<sup>(1)</sup>** |
| Paramedic | 84 (51,5) | 24 (14,7) | 55 (33,7) | 163 (100) |  |
| Hygiene | 16 (80,0) | 2 (10,0) | 2 (10,0) | 20 (100) |  |
| **Percutaneous exposure** |  |  |  |  |  |
| Yes | 80 (52,3) | 8 (12,5) | 21 (32,8) | 153 (100) | 0,943<sup>(2)</sup> |
| No | 35 (54,7) | 21 (13,7) | 52 (34) | 64 (100) |  |

<sup>(1)</sup>: Fisher exact test ; <sup>(2)</sup>: Chi<sup>2</sup> test

Hepatitis B vaccination was associated with marital status and single were less likely to be vaccinated than their counterparts (*p* - value = 0.041). Moreover, compared to physicians, other health professionals were less likely to be vaccinated (OR = 2,23; *p* - value = 0.05) (Table VI). The high cost of vaccine was the most reported reason for non - compliance with hepatitis B vaccination (39 %) (Figure 2).

**Table VI.** Multivariate analysis of factors associated with non – compliance to hepatitis B vaccination among healthcare workers in Yaounde District Hospitals, April (*n* = 217)

| Determinants | aOR | 95 % CI limits | | $p$ - value |
| --- | --- | --- | --- | --- |
|  |  | Lower | Upper |  |
| Male/Female | 1.25 | 0.62 | 2.54 |  |
| Single/Married | 1.82 | 1.03 | 3.23 | **0.041** |
| Year of study: < 12 ans/≥ 12 ans | 1.40 | 0.63 | 3.14 |  |
| Others/Physician | 2.23 | 1.00 | 4.98 | 0.050 |
| Civil servant/Contract worker | 1.14 | 0.63 | 2.07 |  |
| Training on infection control and prevention: No/Yes | 1.05 | 0.59 | 1.87 |  |

aOR: adjusted Odds Ratio

**Figure 2:**
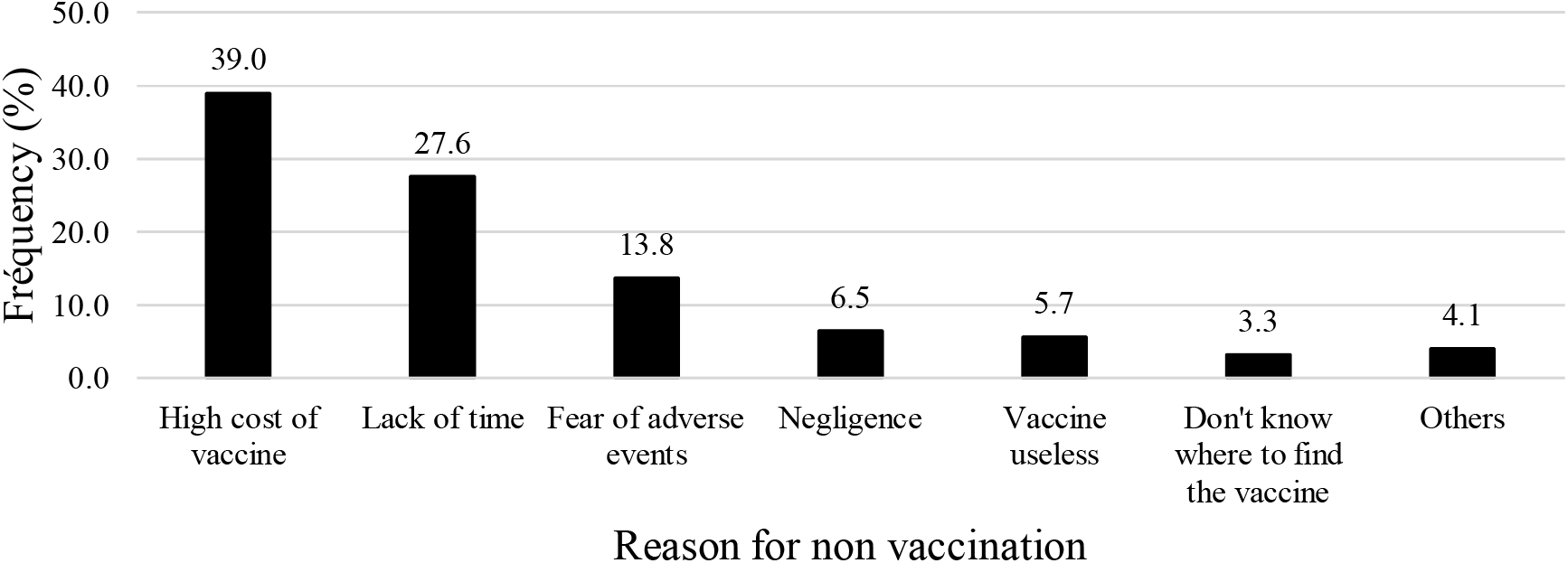
Reasons for non - vaccination against hepatitis B virus in Yaounde District Hospitals, April 2022 (*n =* 115)

## Discussion

The present study outlined the fact, more than half of HCW had experienced an AEB during the last 12 months. This prevalence was relatively lower than that obtained at the Yaounde University Teaching Hospital (YUTH) [23]. Similarly, high levels of AEB were reported in other setting in the Fako Division District hospitals [24] and Tubah DH [25].

Almost one third of our participants had experienced percutaneous exposure to body fluids. Such observations corroborated findings in YUTH [23], elsewhere in Africa [25,26] and in a global meta - analysis [28].

Over their career, more than two third of HCW had experienced AEB. This result was close to findings in similar contexts in Africa [25,16]. Percutaneous exposures varied relatively according to the socioeconomic status of countries. Indeed, lowest prevalence were found in developed countries and highest in developing countries such as Cameroon [28].

The high variability of percutaneous exposure prevalence among health facilities of the same level of care suggests that corrective measures and possible resources should be allocated taking into account this variability for better efficiency.

Men were more affected by occupational injuries compared to women. This could be explain by the fact, men tend to take more risk than female and this negatively impacts their health status[29]. Similar results were found in Ethiopia [10]. In contrast, women were most affected in studies conducted in Laos [13] and Kenya [30]. Contextual factors specific to the different countries or to the study population could explain these differences, sex being considered as a risk factor in various studies [10,11,13,30].

HCW aged between 25 to 49 years were the most exposed to body fluid. This age group is most affected by HIV and hepatitis B as well. This arise the impact of AEB on these infections transmission among HCW [31].

Nearly half of healthcare workers who experience AEB were nurses. Due to the nature of their work, nurses spend more hours in direct contact with patients and undertake more procedures with needles or other instruments that cause percutaneous injuries. They are also called upon to work under pressure because they are at the forefront in the management of emergency cases in the DH healthcare organisation system. nurse This result was similar to those obtained in a global metanalysis [27], Ethiopia [10] and Indonesia [32]. It was higher than those obtained in China [33] and Kenya [30]. China being a developed country, the improvement of the standards of care could explain this low proportion of nurses affected by AEB compared to our study. Nurses remain nevertheless, one of the most at risk occupational groups of percutaneous exposure to blood [34].

Students were almost equally exposed than nurses which may be due to their lack of experience, anterior practical trainings and awareness of the hazards of sharp injuries at that stage of their clinical experience [33]. The risk of infection among that group lies in the fact that preventive vaccinations (Hepatitis B, Tuberculosis) are not systematically required in training institutions during admission.

Although there was a significant difference between the inter - institutional exposure levels, this difference was correlated with the lack of training. On this point, Nkoldongo DH which presented the highest prevalence of AEB was among health facilities with highest proportion of HCW with no past training on infection control and prevention.

Other studies suggest that specific safety precautions and basic infection prevention training in hospitals can improve the operational safety of HCW, thus reducing the occurrence of sharp injuries [35]. A considerable impact of training has been pointed in a European study in preventing AEB and its economic burden among HCW [36]. Public health professionals can provide educational activities to workers with the aim of improving the knowledge and skills necessary to deal with this problem by different methods such as seminars, informative educational boards, pamphlets, and workshops [33,37].

The surgery department had recorded the most cases of percutaneous exposure to blood. The often - urgent nature of the care administered in these departments and the stress that can result could explain the frequent occurrence of accidental occupational injuries. This result was in agreement with those obtained in YUTH [38], Indonesia (31.3 %) [32] and Ethiopia [10] which found, surgical department workers were the most affected by percutaneous AEB.

HCW with less than three years professional experience were the most affected in this study. The lack of skills and training could explain the more frequent occurrence of percutaneous exposure in this occupational group. In this regard, several studies have identified professional experience of less than 5 years as one of the risk factors for percutaneous exposure to blood [17,59,83].

The proportion of workers fully immunized against viral hepatitis B was less than 50 % and varied across hospitals. Moreover, a low proportion of participants were partially vaccinated reflecting the fact they started but did not complete the vaccination process. All this increases the risk and the burden of this disease that are nevertheless preventable [40]. However, this low proportion of fully vaccinated HCW was higher than that obtained at the Bamenda DH (13.9 %) [41] and lower than those obtained by some authors in Kenya [30] and in Serbia [42].

The proportion of fully vaccinated staff was significantly higher among medical staff compared to other occupational groups. Other authors have found similar results [42,43]. Indeed, the financial accessibility of the vaccine for medical personnel, their level of knowledge and perception of occupational risks related to HBV could explain these findings. In this regard, establishing mandatory vaccination against hepatitis B for public health workers could be a possible solution. In France, vaccination against HBV is governed by a law of January 18, 1991 of the Public Health Code which stipulates that any person exercising a professional activity in an establishment or a public or private prevention or care organization exposing him to risks of contamination, must be vaccinated against hepatitis. The excellent vaccination coverage of healthcare personnel against hepatitis B has led to the virtual disappearance of the risk of occupational hepatitis B in developed countries [44].

In addition, other possible solutions include training including items related to risk associated with needle stick and sharp injuries, sensitizations emphasising on the fact hepatitis B vaccination adverse events are limited to common vaccination reactions such injection site pain and redness which are self-limiting most of the time [45,46].

More than a third of healthcare personnel cited the high cost of the vaccine as the main reason for non - compliance to vaccination against HBV. Almost similar results were found at the Bamenda DH [41] and elsewhere in Africa [47].

Single health workers had a greater risk of being non - compliant to vaccination against HBV compared to married workers. Some authors have found in this regard that gender, level of study, grade and the fact of having manuals on biosafety measures at work were significantly associated with vaccination against HBV [42,48,49].

Nearly two thirds of participants who had undergone percutaneous exposure were not fully immunized against HBV. This proportion was high and worrying because in Cameroon, the seroprevalence of HBV in 2017 was high (11.2 %) [39]. So, these exposures could potentially lead to a significant proportion of seroconversion to viral hepatitis B among HCW. A study in YUTH had found that one third of percutaneous exposure victims were non - vaccinated against hepatitis B [23] ; this is probably due to the retrospection of only 3 months of the cases of AEB identified, applied in this study against 12 months for ours.

## Conclusion

Healthcare workers of Yaounde DH reported a high prevalence of AEB. This reflects high risk of a healthcare related infections exposure among HCW. Vaccination coverage against hepatitis B among HCW were low indicating high exposure to HBV. There is an urgent need to implement AEB prevention strategies and strengthen the observance of standard precaution measures including preventive vaccination targeting hepatitis B in DH at the assumption of duties by clinicians.

## Data Availability

All data produced in the present study are available upon reasonable request to the authors

## Declaration

### Ethical Approval Statement

The protocol was approved by the Regional Human Health Committee of the Centre (CRERSH - Ce) with the ethical clearance: CE N° 2245/CRERSHC/2021. An authorisation was obtained from all District Medical Officer. Participants’ informed consent was obtained prior to data collection.

## Acknowledgments

Our gratitude goes to healthcare workers who agreed to participate in this study.

## Footnotes

### Contributors

FZLC— investigation, methodology, data curation and analysis, resources, visualisation, writing original draft (review and editing). IT— conceptualisation, methodology, data analysis, validation, roles/writing (original draft); writing (review and editing), EEL—writing (review and editing). HGK—writing (review and editing), F-XM-K —writing (review and editing).

## References

1. World Health Organization. Aide-memoire for a strategy to protect health workers from infection with bloodborne viruses [Internet]. World Health Organization; 2003 [cited 2021 Sep 8]. Report No.: WHO/BCT/03.11. Available from: https://apps.who.int/iris/handle/10665/68354

2. Annette PU, Elisabetta R, Yvan H. Estimation of the global burden of disease attributable to contaminated sharps injuries among health-care workers. Am J Ind Med [Internet]. 2005 Dec [cited 2021 Sep 8];48(6). Available from: https://pubmed.ncbi.nlm.nih.gov/16299710/

3. Westermann C, Peters C, Lisiak B, Lamberti M, Nienhaus A. The prevalence of hepatitis C among healthcare workers: a systematic review and meta-analysis. Occup Environ Med. 2015 Dec;72(12):880–8.

4. Em B, It W, Cn S, Me C. Risk and management of blood-borne infections in health care workers. Clin Microbiol Rev [Internet]. 2000 Jul [cited 2021 Sep 8];13(3). Available from: https://pubmed.ncbi.nlm.nih.gov/10885983/

5. Himmelreich H, Rabenau HF, Rindermann M, Stephan C, Bickel M, Marzi I, et al. The management of needlestick injuries. Dtsch Arzteblatt Int. 2013 Feb;110(5):61–7.

6. World Health Organization (WHO). Needlestick injuries [Internet]. 2019. Available from: https://www.who.int/occupational_health/topics/needinjuries/en/.

7. Auta A, Adewuyi EO, Tor-Anyiin A, Edor JP, Kureh GT, Khanal V, et al. Global prevalence of percutaneous injuries among healthcare workers: a systematic review and meta-analysis. Int J Epidemiol. 2018 Dec 1;47(6):1972–80.

8. Domkam IK, Sonela N, Kamgaing N, Takam PS, Gwom LC, Betilene TMA, et al. Prevalence and risk factors to HIV-infection amongst health care workers within public and private health facilities in Cameroon. Pan Afr Med J. 2018;29:158.

9. Mengistu DA, Tolera ST. Prevalence of occupational exposure to needlestick injury and associated factors among healthcare workers of developing countries: Systematic review. J Occup Health. 2020 Dec 12;62(1):e12179.

10. Assen S, Wubshet M, Kifle M, Wubayehu T, Aregawi BG. Magnitude and associated factors of needle stick and sharps injuries among health care workers in Dessie City Hospitals, north east Ethiopia. BMC Nurs. 2020 Apr 21;19(1):31.

11. Hassanipour S, Sepandi M, Tavakkol R, Jabbari M, Rabiei H, Malakoutikhah M, et al. Epidemiology and risk factors of needlestick injuries among healthcare workers in Iran: a systematic reviews and meta-analysis. Environ Health Prev Med. 2021 Apr 1;26(1):43.

12. Bekele T, Gebremariam A, Kaso M, Ahmed K. Factors Associated with Occupational Needle Stick and Sharps Injuries among Hospital Healthcare Workers in Bale Zone, Southeast Ethiopia. PLOS ONE. 2015 Oct 15;10(10):e0140382.

13. Matsubara C, Sakisaka K, Sychareun V, Phensavanh A, Ali M. Prevalence and risk factors of needle stick and sharp injury among tertiary hospital workers, Vientiane, Lao PDR. J Occup Health. 2017;59(6):581–5.

14. Goel V, Kumar D, Lingaiah R, Singh S. Occurrence of Needlestick and Injuries among Healthcare Workers of a Tertiary Care Teaching Hospital in North India. J Lab Physicians. 2017 Mar;9(1):20–5.

15. Mémoire infirmierL: La gestion des accidents d’exposition au sang chez le personnel paramédical [Internet]. ParaMedical. 2020 [cited 2021 Oct 7]. Available from: https://paramedz.com/infirmier-memoire/memoire-infirmier-la-gestion-des-accidents-dexposition-au-sang-chez-le-personnel-paramedical/

16. Ziraba AK, Bwogi J, Namale A, Wainaina CW, Mayanja-Kizza H. Sero-prevalence and risk factors for hepatitis B virus infection among health care workers in a tertiary hospital in Uganda. BMC Infect Dis. 2010 Dec;10(1):191.

17. Mossburg S, Agore A, Nkimbeng M, Commodore-Mensah Y. Occupational Hazards among Healthcare Workers in Africa: A Systematic Review. Ann Glob Health. 2019 Jun 6;85(1):78.

18. Fritzsche C, Becker F, Hemmer CJ, Riebold D, Klammt S, Hufert F, et al. Hepatitis B and C: neglected diseases among health care workers in Cameroon. Trans R Soc Trop Med Hyg. 2013 Mar;107(3):158–64.

19. Mueller A, Stoetter L, Kalluvya S, Stich A, Majinge C, Weissbrich B, et al. Prevalence of hepatitis B virus infection among health care workers in a tertiary hospital in Tanzania. BMC Infect Dis. 2015 Sep 23;15:386.

20. Kong SYJ, Wi DH, Ro YS, Shin SD, Jeong J, Kim YJ, et al. Changes in the healthcare utilization after establishment of emergency centre in Yaoundé, Cameroon: A before and after cross-sectional survey analysis. PLOS ONE. 2019 Feb 8;14(2):e0211777.

21. Bonny A, Tibazarwa K, Mbouh S, Wa J, Fonga R, Saka C, et al. Epidemiology of sudden cardiac death in Cameroon: the first population-based cohort survey in sub-Saharan Africa. Int J Epidemiol. 2017 Aug 1;46(4):1230–8.

22. Reports | DHIS2 [Internet]. [cited 2022 Oct 15]. Available from: https://dhis-minsante-cm.org/dhis-web-reports/index.html#/data-set-report

23. Nouetchognou JS, Ateudjieu J, Jemea B, Mbanya D. Accidental exposures to blood and body fluids among health care workers in a Referral Hospital of Cameroon. BMC Res Notes. 2016 Dec;9(1):94.

24. Ngwa CH, Ngoh EA, Cumber SN. Assessment of the knowledge, attitude and practice of health care workers in Fako Division on post exposure prophylaxis to blood borne viruses: a hospital based cross-sectional study. Pan Afr Med J. 2018 Oct 12;31:108.

25. Aminde LN, Takah NF, Dzudie A, Bonko NM, Awungafac G, Teno D, et al. Occupational Post-Exposure Prophylaxis (PEP) against Human Immunodeficiency Virus (HIV) Infection in a Health District in Cameroon: Assessment of the Knowledge and Practices of Nurses. PLOS ONE. 2015 Apr 16;10(4):e0124416.

26. Auta A, Adewuyi EO, Tor-Anyiin A, Aziz D, Ogbole E, Ogbonna BO, et al. Health-care workers’ occupational exposures to body fluids in 21 countries in Africa: systematic review and meta-analysis. Bull World Health Organ. 2017 Dec 1;95(12):831–841F.

27. Bouya S, Balouchi A, Rafiemanesh H, Amirshahi M, Dastres M, Moghadam MP, et al. Global Prevalence and Device Related Causes of Needle Stick Injuries among Health Care Workers: A Systematic Review and Meta-Analysis. Ann Glob Health. 2020 Apr 6;86(1):35.

28. Mengistu DA, Tolera ST, Demmu YM. Worldwide Prevalence of Occupational Exposure to Needle Stick Injury among Healthcare Workers: A Systematic Review and Meta-Analysis. Can J Infect Dis Med Microbiol. 2021 Jan 29;2021:e9019534.

29. Asai T, Matsumoto S, Matsumoto H, Yamamoto K, Shingu K. Prevention of needle-stick injury. Efficacy of a safeguarded intravenous cannula. Anaesthesia. 1999 Mar;54(3):258–61.

30. Mbaisi EM, Ngángá Z, Wanzala P, Omolo J. Prevalence and factors associated with percutaneous injuries and splash exposures among health-care workers in a provincial hospital, Kenya, 2010. Pan Afr Med J [Internet]. 2013 Jan 6 [cited 2021 Sep 6];14(10). Available from: https://www.panafrican-med-journal.com/content/article/14/10/full

31. Singhal V, Bora D, Singh S. Hepatitis B in health care workers: Indian scenario. J Lab Physicians [Internet]. 2009 Jul [cited 2021 Sep 8];1(2). Available from: https://pubmed.ncbi.nlm.nih.gov/21938248/

32. Yunihastuti E, Ratih DM, Aisyah MR, Hidayah AJ, Widhani A, Sulaiman AS, et al. Needlestick and sharps injuries in an Indonesian tertiary teaching hospital from 2014 to 2017: a cohort study. BMJ Open. 2020 Dec 1;10(12):e041494.

33. Lin J, Gao X, Cui Y, Sun W, Shen Y, Shi Q, et al. A survey of sharps injuries and occupational infections among healthcare workers in Shanghai. Ann Transl Med. 2019 Nov;7(22):678.

34. Kaweti G, Abegaz T. Prevalence of percutaneous injuries and associated factors among health care workers in Hawassa referral and adare District hospitals, Hawassa, Ethiopia, January 2014. BMC Public Health. 2016 Jan 5;16(1):8.

35. Dilie A, Amare D, Gualu T. Occupational Exposure to Needle Stick and Sharp Injuries and Associated Factors among Health Care Workers in Awi Zone, Amhara Regional State, Northwest Ethiopia, 2016. J Environ Public Health. 2017 Aug 10;2017:e2438713.

36. Aziz AM. Do training and needle-safety devices prevent needlestick injuries? A systematised review of the literature. Br J Nurs Mark Allen Publ. 2018 Sep 6;27(16):944–52.

37. Afridi A, Kumar A, Sayani R. Needle Stick Injuries – Risk and Preventive Factors: A Study among Health Care Workers in Tertiary Care Hospitals in Pakistan. Glob J Health Sci. 2013 Apr 14;5(4):p85.

38. Mbanya D, Ateudjieu J, Tagny CT, Moudourou S, Lobe MM, Kaptue L. Risk Factors for Transmission of HIV in a Hospital Environment of Yaoundé, Cameroon. Int J Environ Res Public Health. 2010 May;7(5):2085–100.

39. Bazie GW. Factors Associated with Needle Stick and Sharp Injuries Among Healthcare Workers in North East Ethiopia. Risk Manag Healthc Policy. 2020 Nov 3;13:2449–56.

40. Pattyn J, Hendrickx G, Vorsters A, Van Damme P. Hepatitis B Vaccines. J Infect Dis. 2021 Sep 30;224(12 Suppl 2):S343–51.

41. Ngum AM, Laure SJ, Tchetnya X, Tambe TA, Ngwayu CN, Wirsiy FS, et al. Vaccination against Hepatitis B among health care workers in the Bamenda Health District: influence of knowledge and attitudes, Cameroon. Pan Afr Med J. 2021 Dec 9;40:216.

42. Markovic-Denic L, Maksimovic N, Marusic V, Vucicevic J, Ostric I, Djuric D. Occupational Exposure to Blood and Body Fluids among Health-Care Workers in Serbia. Med Princ Pract. 2015;24(1):36–41.

43. Ndongo CB, Eteki L, Siedner M, Mbaye R, Chen J, Ntone R, et al. Prevalence and vaccination coverage of Hepatitis B among healthcare workers in Cameroon: A national seroprevalence survey. J Viral Hepat. 2018 Dec;25(12):1582–7.

44. Laraqui O, Laraqui S, Tripodi D, Zahraoui M, Caubet A, Christian V, et al. Évaluation des connaissances, attitudes et pratiques sur les accidents d’exposition au sang en milieu de soins au Maroc. Med Mal Infect - MED MAL INFEC. 2008 Dec 1;38:658–66.

45. Safety Information for Hepatitis B Vaccines | Vaccine Safety | CDC [Internet]. 2022 [cited 2023 Mar 5]. Available from: https://www.cdc.gov/vaccinesafety/vaccines/hepatitis-b-vaccine.html

46. Liu Y, Zhang M, Yang M, Chen Q. Adverse Events of Vaccination against Hepatitis B Virus in Post-Marketing Surveillance from 2005 to 2017 in Guangdong Province, China. Vaccines. 2022 Jul 6;10(7):1087.

47. Shah SM, Rodin H, Pogemiller H, Magbagbeola O, Ssebambulidde K, Zewde A, et al. Hepatitis B Awareness and Vaccination Patterns among Healthcare Workers in Africa. Am J Trop Med Hyg. 2020 Dec;103(6):2460–8.

48. Al-Abhar N, Moghram GS, Al-Gunaid EA, Al Serouri A, Khader Y. Occupational Exposure to Needle Stick Injuries and Hepatitis B Vaccination Coverage Among Clinical Laboratory Staff in Sana’a, Yemen: Cross-Sectional Study. JMIR Public Health Surveill. 2020 Mar 31;6(1):e15812.

49. Bhattarai S, Kc S, Pradhan PM, Lama S, Rijal S. Hepatitis B vaccination status and Needle-stick and Sharps-related Injuries among medical school students in Nepal: a cross-sectional study. BMC Res Notes. 2014 Nov 3;7(1):774.

